# Disruption of the renal service during COVID-19 pandemic, physicians survey responses from 5 countries

**DOI:** 10.1101/2020.06.30.20143354

**Authors:** Tarek Samy Abdelaziz, Eman Elsayed, Moataz Fatthy

**Author notes:** Corresponding Tarek Samy Abdelaziz, M.D., Department of Internal medicine-Renal Division- Kasr Alainy Hospitals- Cairo University- Egypt.

## Abstract

The outbreak of COVID-19 has significant impact on health care systems. The workflow in renal care facilities and dialysis facilities is complex. The aim of his study was to explore multinational renal physician perspective about the disruption of outflow of renal service amid COVID-19 pandemic.

**Methods:** we distributed a questionnaire electronically to renal physicians of various grades from 5 countries. The questionnaire was formulated of 9 questions with domains centered on the acute and chronic renal services

**Results:** 97 physicians took part in the survey from 5 countries. 58% of the participating renal physicians agreed that there has been disruption to the acute service offered to renal patients. 38% of the physicians who took the survey report disruption to the hemodialysis facilities.

**Conclusion:** At this challenging time of the pandemic, it is important to perform audits and quality control to explore the deficiencies of acute and chronic renal care pathways.

## Introduction

Since the outbreak of COVID-19 pandemic, caused by SARS-CoV-2, the healthcare systems around the world have seen one of the biggest challenges. Drastic measures have been taken by the health authorities to fight and mitigate the pandemic. The challenges to both acute and chronic renal care services are paramount (1). We aimed to reach out to renal physicians of variable grades to explore their experience about the outflow of both acute and chronic renal care pathways.

## Methods

We developed a survey and distributed it to health-care professionals in 5 countries around the world (Egypt, Italy, Turkey, Spain and India). The survey consisted of 9 questions with two main domains. Some questions were addressing the adequacy and timely application of acute renal care to patients amid the COVID-19 pandemic; the other questions were about dialysis units’ preparedness. Renal physicians were invited to take part through email. The questions are shown in table (1)

**Table 1.**
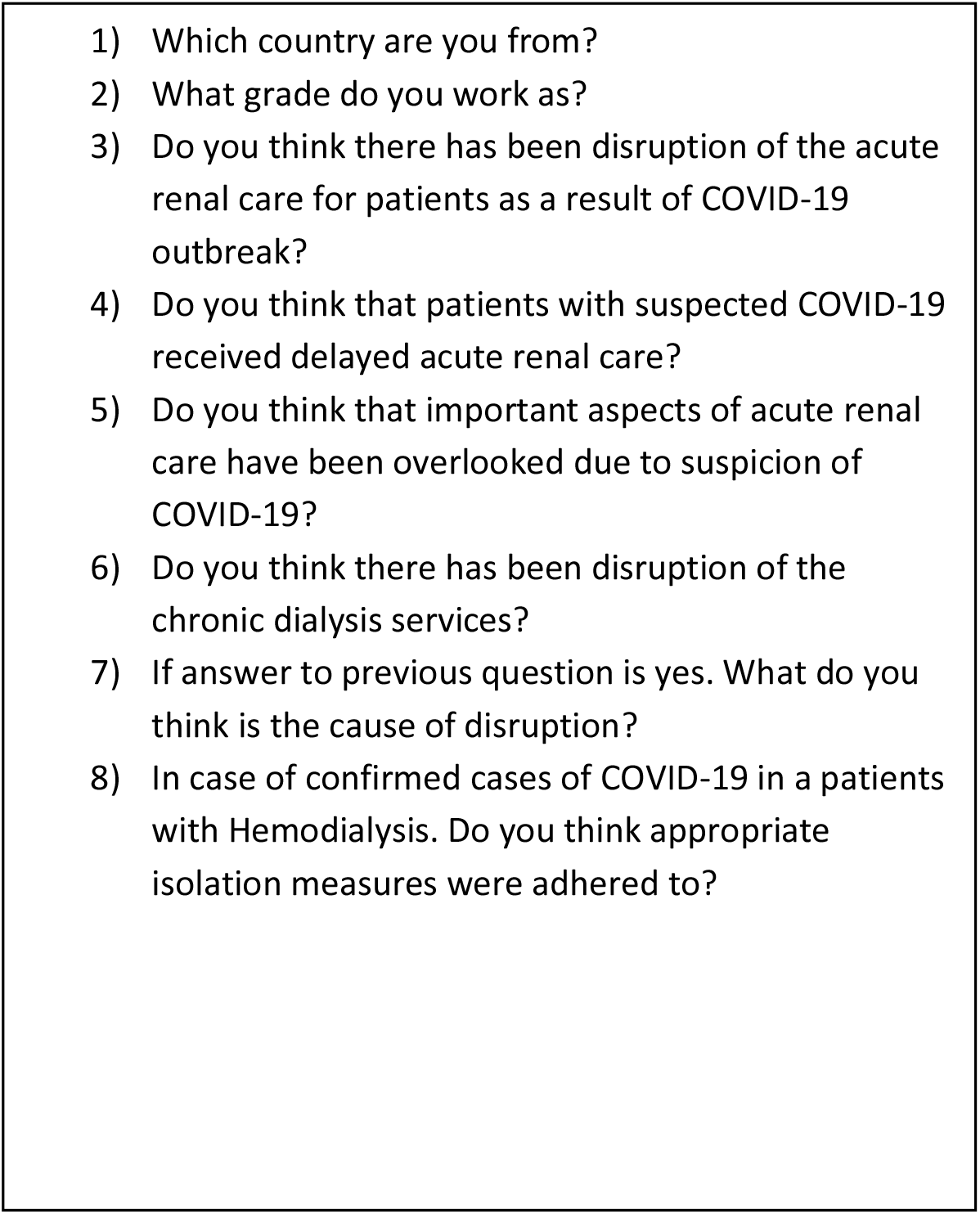
Survey of assessment of Renal service outflow during COVID-19 pandemic

## Results

97 physicians took part in the survey from 5 countries. The countries represented in this survey are Egypt, Italy, Turkey, Spain and India. 75% of the participants work in tertiary care facilities, the remaining 25 % work in secondary care facilities.23% of the participants was consultants; 65% were trainee and 12% were managers of renal service.

### Responses

#### Disruption of the acute renal care

58% of the participating renal physicians agreed that there has been disruption to the acute service offered to renal patients. Suspected cases of SARS-CoV2 virus experienced a varied degree of delay in initiating acute care pathways. 63 percent of the physicians report experiencing temporary disruption of acute hemodialysis units since the outbreak. The disruption of acute hemodialysis was attributed to shortage of medical staff due to sick leaves or due to the presence of suspected or confirmed cases of COVID-19. 25% of physicians agree that it has been challenging to establish appropriate and timely diagnosis of renal diseases, in the acute settings, due to conflict between acute renal care pathway and COVID-19 pathway.

#### Disruption to the chronic hemodialysis service

38% of the physicians who took the survey report disruption to the hemodialysis facilities. The disruption was caused by a number of factors. Accommodation of incipient dialysis patients, staff absenteeism on sick leaves or the diagnosis of new cases of COVID-19. 43% agreed that appropriate isolation measures were adhered to.

### Admission policies

Admission policies to acute healthcare facilities were variable and in some occasions interfered with timely management of patients who needed acute or chronic renal care and, were suspected of having COVID-19. The delay was due to the requirement of obtaining two negative SARS-CoV-2 nucleic acid tests for patients with criteria suspicious of COVID-19

## Discussion

In our survey, we explored the experience of renal physicians from 5 different countries. The core of survey was the effect of COVID-19 on the provision of acute and chronic renal service. 58% of the participants agreed that there has been disruption of the acute renal pathway. There have been different guidelines addressing diffent domains of renal care amid COVID-19. Previous reports have highlighted the importance of optimizing resources at the time of increased demand (1). Admission policies of suspected or confirmed cases of COVID-19 were different form one country to another, as reported by renal physicians in this survey. In Wuhan suspected cases, among hemodialysis patients, were required to have 2 negative nucleic acid tests before being admitted to the dialysis facilities (2).

## Conclusion

Renal physicians from 5 countries report temporary disruption of the acute and chronic hemodialysis facilities. At this challenging time of the pandemic, it is important to perform audits and quality control to explore the deficiencies of acute and chronic renal care pathways.

## Data Availability

available upon request

## Acknowledgment

We are grateful to all renal physicians who responded to the survey.

## References

1. Burgner A, Ikizler TA, Dwyer JP. COVID-19 and the inpatient dialysis unit. CLIN J AM SOC NEPHROL 15(5): 720, 2020

2. Li J, & Xu G. Lessons from the experience in wuhan to reduce risk of COVID-19 infection in patients undergoing long-term hemodialysis. CLIN J AM SOC NEPHROL 15(5): 717, 2020

